# High-resolution vector-borne disease infection risk mapping with Area-to-Point Poisson Kriging and Species Distribution Modeling

**DOI:** 10.1101/2024.05.03.24306806

**Authors:** Szymon Moliński

**Affiliations:** DataverseLabs, Wrocław, Poland

**Keywords:** Species distribution modeling, Ticks, Lyme Disease, Ixodes, Poisson Kriging, Area-to-Point Kriging, Population at risk

## Abstract

**Background:** Disease infection data is usually aggregated and shared as a sum of infections in a given area over time. This data is presented as choropleth maps. The aggregation process protects privacy and simplifies decision-making but introduces visual bias for large areas and sparsely populated places. Moreover, aggregated areas of varying sizes cannot be simply used as the input for complex ecological models, which are based on data retrieved at higher resolution on regular grids. The issue is especially painful for vector-borne diseases, e.g. Lyme Disease, where infection risk is closely related to vector species and their ecological niche.

**Methods:** The paper presents the method of obtaining high-resolution risk maps using a pipeline with two components: (1) spatial disaggregation component, which transforms incidence rate aggregates into the point-support model using Area-to-Point Poisson Kriging, and (2) species distribution modeling component, which detects areas where ticks bite is more likely using MaxEnt model. The first component disaggregates Lyme Disease incidence rates summed over counties in Poland, Central Europe, in 2015. The second component uses ticks occurrence maps, Leaf Area Index, Normalized Difference Vegetation Index, Land Surface Temperature derived from Earth Observation satellites, and Digital Elevation Model.

The final weighted population-at-risk map is a product of both components outputs. The pipeline is built upon open source and open science projects, and it is reusable.

**Results:** The presented pipeline creates high-resolution risk maps: vector occurrence probability map, population-at-risk map, and weighted population-at-risk map which includes information about local infections and about vector species. The final maps have much better resolution than aggregated incidence rates. Visual bias for population-at-risk maps is removed, and unpopulated areas are not presented on the map.

**Conclusions:** The pipeline might be used for other vector-borne diseases. The final weighted population-at-risk map might be used as an input for another analytical model requiring high-resolution data placed over a regular grid. The pipeline removes visual bias and transforms aggregated data into a high-resolution point-support layer.

## Background

Lyme Disease incidence rates collected worldwide are aggregated over administrative units. Spatial aggregation protects patients’ privacy and simplifies decision-making by governance bodies. However, the aggregation process has serious downsides. Administrative units have varying shapes and sizes (i). Larger areas on choropleth maps tend to get more attention from human viewers, and perceptual homogeneity of incidence rates across the area is misleading - especially for sparsely populated regions [1].

Size and extent of aggregated incidence rates are not comparable to other spatial data sources (ii) used for infection risk modeling. The examples are biotic and abiotic variables available at a higher resolution that correlate with ticks’ abundance and questing behavior.

Local weather is one of those factors. Temperature and humidity directly influence ticks activity [2, 3]. Climate and climate change variables are other variables worth considering. Milder winters and higher temperatures increase tick distribution worldwide. In Europe, ticks are sampled at higher altitudes and latitudes [4]. The number of ticks increases in the northern United States and Canada [5]. Land cover features are the next group of covariates indirectly informing about ticks’ possible range [6, 7, 8]. Finally, there are multiple biotic factors affecting tick abundance and infection risk: vegetation type [9], host availability [10], and host behavior [11].

Using this data, creating the infection risk model with a fine-scale resolution is possible. The assumption is that the deconvoluted risk infection map might be weighted by the probability of the vector’s occurrence derived from the species distribution model. The first step in the pipeline is the transformation of areal aggregates of incidence rates into high-resolution blocks and using those blocks as input for the spatial infection risk model (deconvolution). The second part is the estimation of the species distribution map. Tick spatial distribution is derived from the climate, spatial features, and vector occurrence samples. The final model is an output from the merged deconvoluted infection risk map and species distribution map. It presents a high-resolution infection risk map, incorporating information about the vector abundance in a given area.

The study presents a methodology for retrieving the risk of vector-borne disease infection at a scale finer than the incidence rates aggregated over administrative regions. The system uses geostatistics, machine learning, remote sensing data, and information about hard-bodied ticks species to create a high-resolution Lyme disease infection risk map for Poland. Associated Python code and data are available in a Zenodo archive [12].

## Methods

### Data Sources

The system uses spatial (GIS), remote sensing, and public health datasets limited to the area of Central Europe (Poland and Germany). Processed datasets and metadata are available in the Zenodo repository.

The datasets used for the population-at-risk estimation are:

- The population density grid in Poland from the Central Statistical Office [13]. The baseline grid size was 1×1 kilometer. Transformed into 5×5 kilometers blocks.
- The incidence rates of Lyme Disease in each Polish county in 2015 (Figure 1). Data was retrieved manually from each voivodship sanitary-epidemiological station.
- Administrative borders of Polish counties.

**Figure 1:**
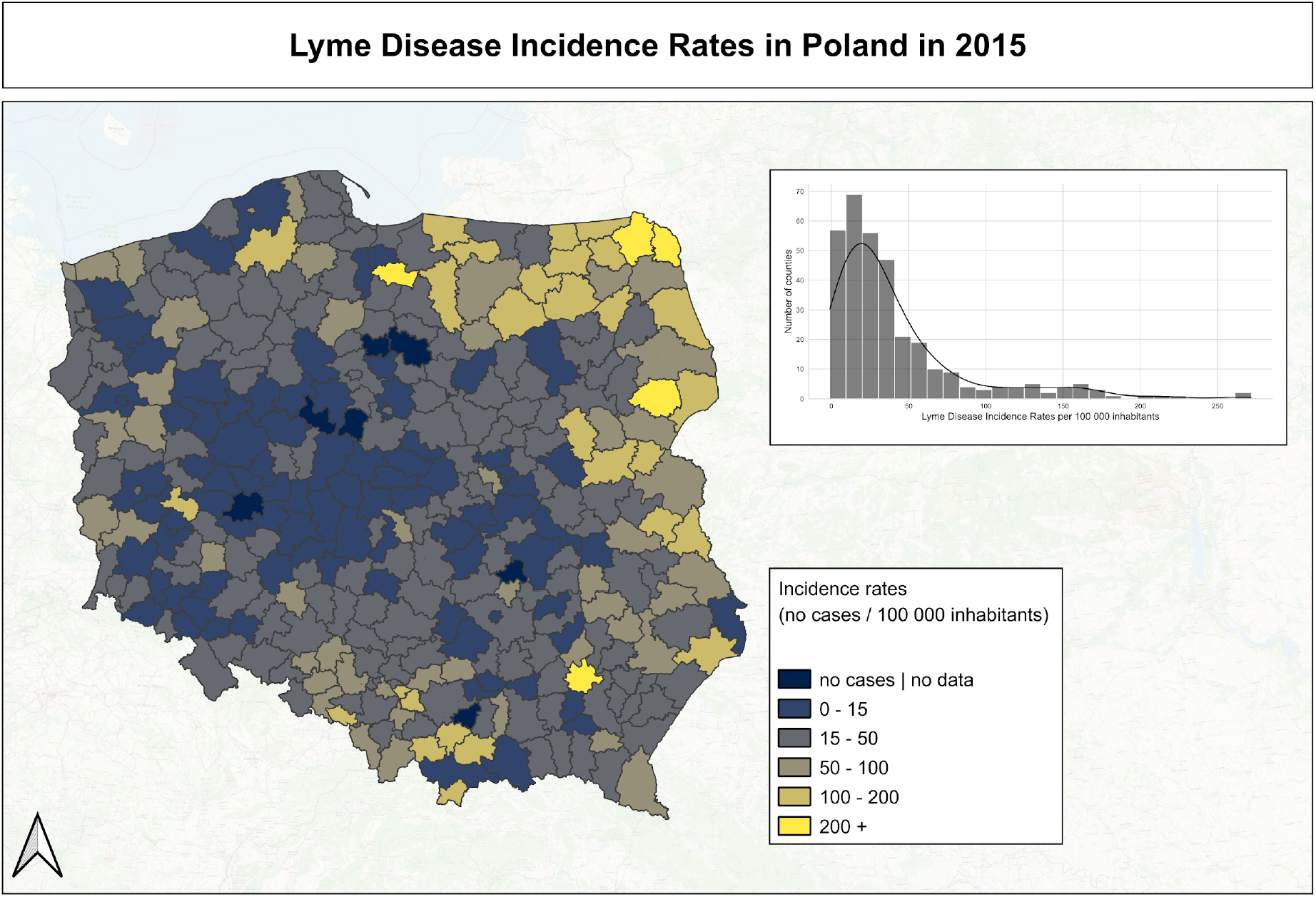
Spatial distribution of Lyme Disease incidence rates in Polish counties.

The ticks distribution model uses:

- Ticks occurrence locations in Germany [14],
- bioclimatic and environmental factors with spatial resolution 1×1 kilometer: Land Surface Temperature, Leaf Area Index, Normalized Difference Vegetation Index, Digital Elevation Model (EU-DEM) for areas of Germany and Poland. Data was obtained from the Copernicus Land Monitoring Service [15].

### Poisson Kriging: spatial interpolation of population at risk

The population-at-risk map was developed using the Poisson Kriging technique. Poisson Kriging was used for smoothing and areal deconvolution in other public health studies related to cancer mortality, cholera, and dysentery [1, 16, 17].

The incidence rate is defined as *z* (*u*_α_) = 10^5^ * *d*(*u*_α_)/*n*(*u*_α_), where *d*(*u*_α_) is the number of cases in a fixed time interval and *n*(*u*_α_) is the total population in a region. The incidence rate is multiplied by 100,000, which indicates the number of cases for 100,000 inhabitants. The number of cases is interpreted as a random variable *D*(*u*_α_) that follows the Poisson distribution with one parameter - the expected number of cases per year. It can be defined as a product of the local risk *r*(*u*_α_) and population density on a given area *n*(*u*_α_). The local risk *r*(*u*_α_) might be estimated as a linear combination of the local incidence rate *z* (*u*_α_) and incidence rates observed along *K* neighboring areas *u*_*i*_

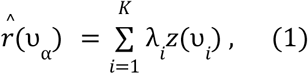

Weights λ_*i*_ assigned to *K* neighbors are obtained after solving the system of linear equations:

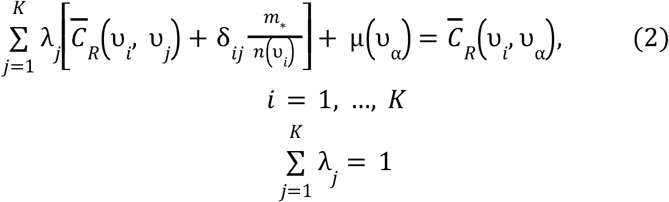

The technique can be used for data where spatial autocorrelation is significant. The semivariogram analysis and modeling precede kriging. The semivariogram measures the dissimilarity between observations in the function of a distance. When semivariances at short distances are low and rise with increasing distance, the process is spatially correlated. (Compare it with Figure 3). Semivariance is the halved mean squared error between point observations and all other points in a specified range (bin).

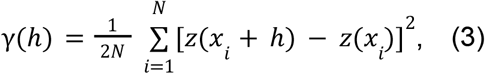

Semivariance analysis, semivariogram modeling, and spatial interpolation steps were performed in the Python package Pyinterpolate [18].

### Vector distribution model

Choropleth map of Lyme Disease incidence rates informs about the medical condition, but it tells nothing about the vector distribution. Thus, the spatial risk may be miscalculated - tick bites occur in different places where infected people live. The full information about the infection risk should include the vector density at a given area or the probability of the vector occurrence. The occurrence probability is obtained from the species distribution model. Models of this type are estimated from the information about bio-climatic factors measured in places where species are sampled. The model analyses the correlation between bioclimatic variables and vector occurrences/absences. The popular approach is to use remote sensing data to study vector species [19].

The maximum entropy MaxEnt model is the most popular species distribution model architecture [20]. The risk-modeling pipeline uses an implementation of MaxEnt from the Elapid Python package [21]. The result of the calculations is a tick occurrence probability map (Figure 4). The pipeline model uses:

- points where Lyme Disease (*D*.*marginatus, D. reticulatus, I. hexagonus, I*.*ricinus, I*.*trianguliceps*) transmitting ticks were sampled, within Germany border [22]
- Leaf Area Index, Normalized Difference Vegetation Index (mean and standard deviation from years 2005-2015), Land Surface Temperature (mean and standard deviation from years 2017-2020), and Digital Elevation Model EU DEM 1.1 from Copernicus Land Monitoring Services [15].

Species Distribution Model input - training set - comes from Germany because tick observations for this country are available, and Germany has a climate and species similar to Poland.

### Complex vector-borne disease infection risk model

Poisson Kriging model derives the population-at-risk of Lyme Disease infection, and the species distribution model generates a probability map of vector occurrence. Both parameters affect local infection risk, and might be related by following reasoning:

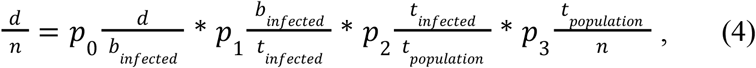

where *d* - number of cases, *n* - population, *b*_*infected*_ - number of bites of infected ticks, *t*_*infected*_ - number of infected ticks, *t*_*population*_ - ticks total population. All variables are valid for a given region and time.Variables *p*_0_, *p*_1_, *p*_2_, *p*_3_ represent unknown weighting factors for each part of the equation. The first part 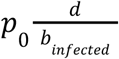 represents the probability of successful disease transmission from vector to host. The second part 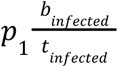 is the probability of being beaten by infected tick, the next part 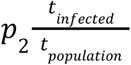 is the probability of being a disease vector, and the last part 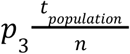 is the probability of spotting tick in the local area. Based on the equation it can be said that the presented approach of merging Poisson Kriging model with species distribution model estimates the last part of the first and the last of the equation (Poisson Kriging represents risk of being infected, and species distribution model links host and vector populations).

Merging both models allows for weighting local population at risk 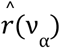 with probability of ticks (vectors) occurrence *P*_*_ (ν_α_). Species-probability weighting lowers local population-at-risk indices when the probability of occurrence is below 0.5, and the risk increases for probabilities greater than 0.5. The equation for normalized risk index is:

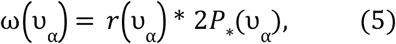

The high-level flow with input datasets and model outputs is presented in the diagram (Figure 2).

**Figure 2:**
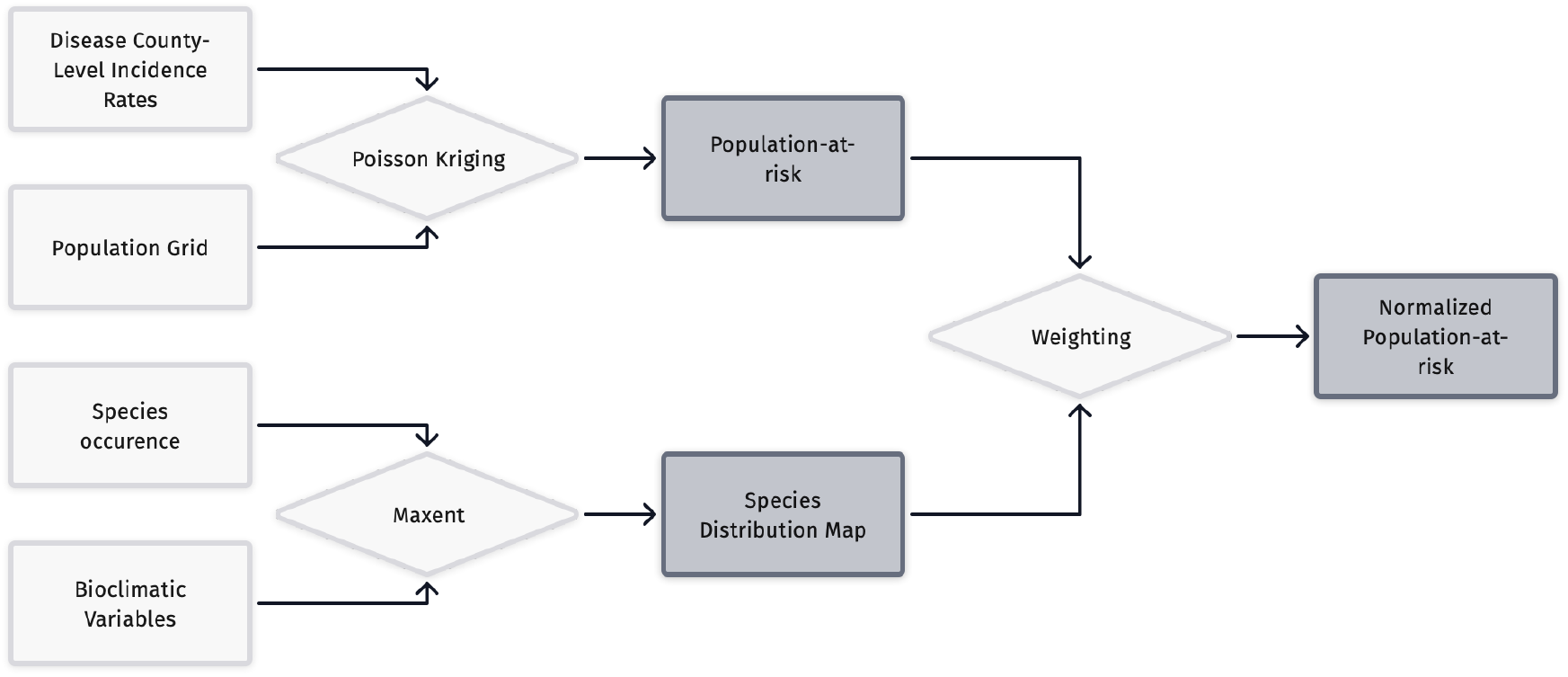
Modeling steps.

## Results

The first step is to check if Lyme Disease incidence rates in neighboring areas are similar. It can be done by analyzing the semivariogram, the plot of squared error between neighboring areas as a function of the distance between them (Figure 3). Each county area was transformed to its centroid, and semivariance was measured between those centroids. The plot shows that neighboring counties have similar incidence rates.

**Figure 3:**
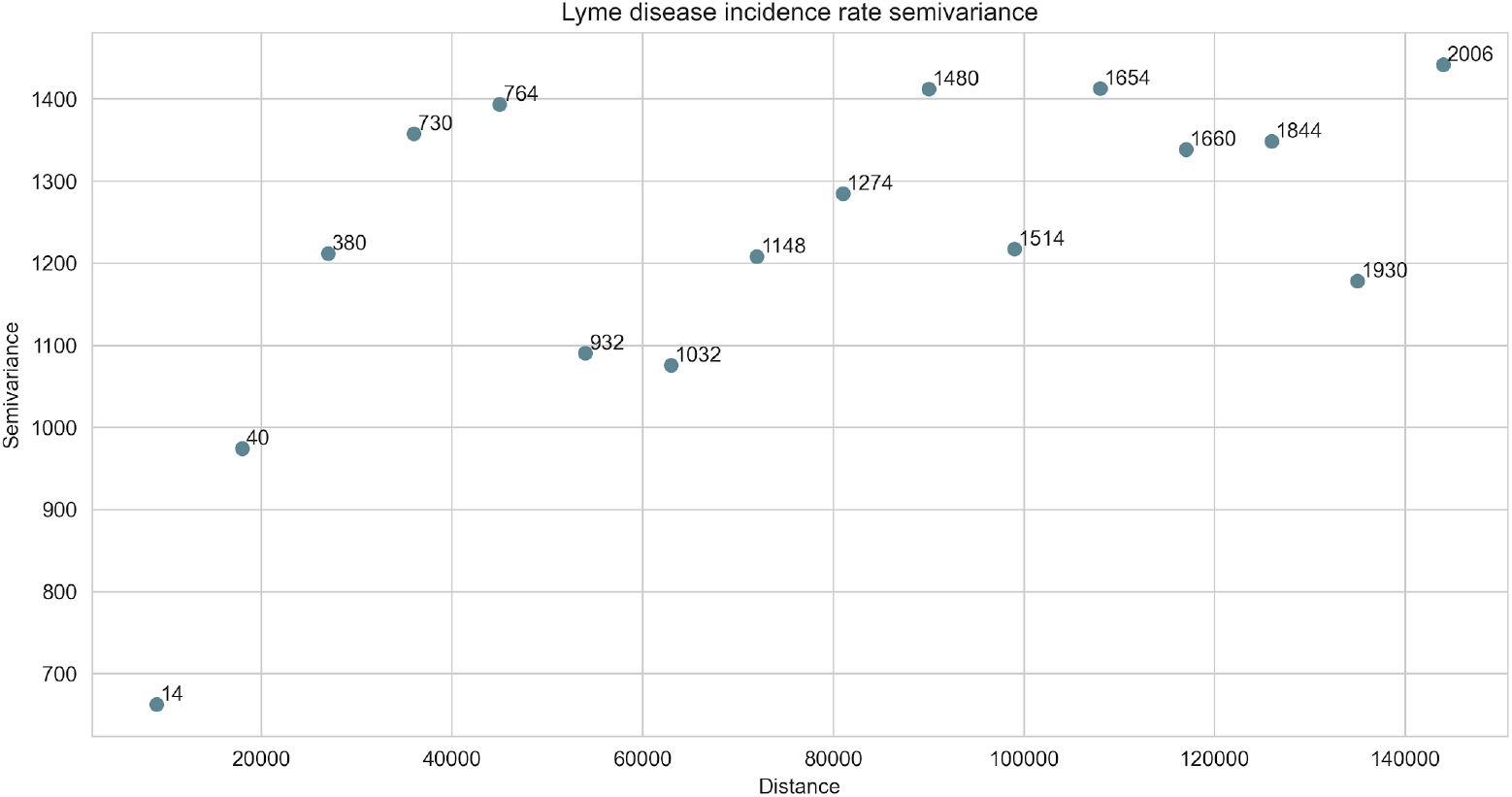
Dissimilarity of point pairs representing county centroids over a rising distance. Numbers show how many point pairs are averaged to obtain a result.

Semivariances increase up to 60 kilometers, then oscillate around the maximum variance threshold. This means that the spatial correlation between neighboring areas is noticeable up to 60 kilometers, thus the Poisson Kriging model’s maximum range is set to this distance. The input for the population-at-risk are Lyme Disease incidence rates in Polish counties (Figure 1) and the Population Density grid (Figure 4).

**Figure 4:**
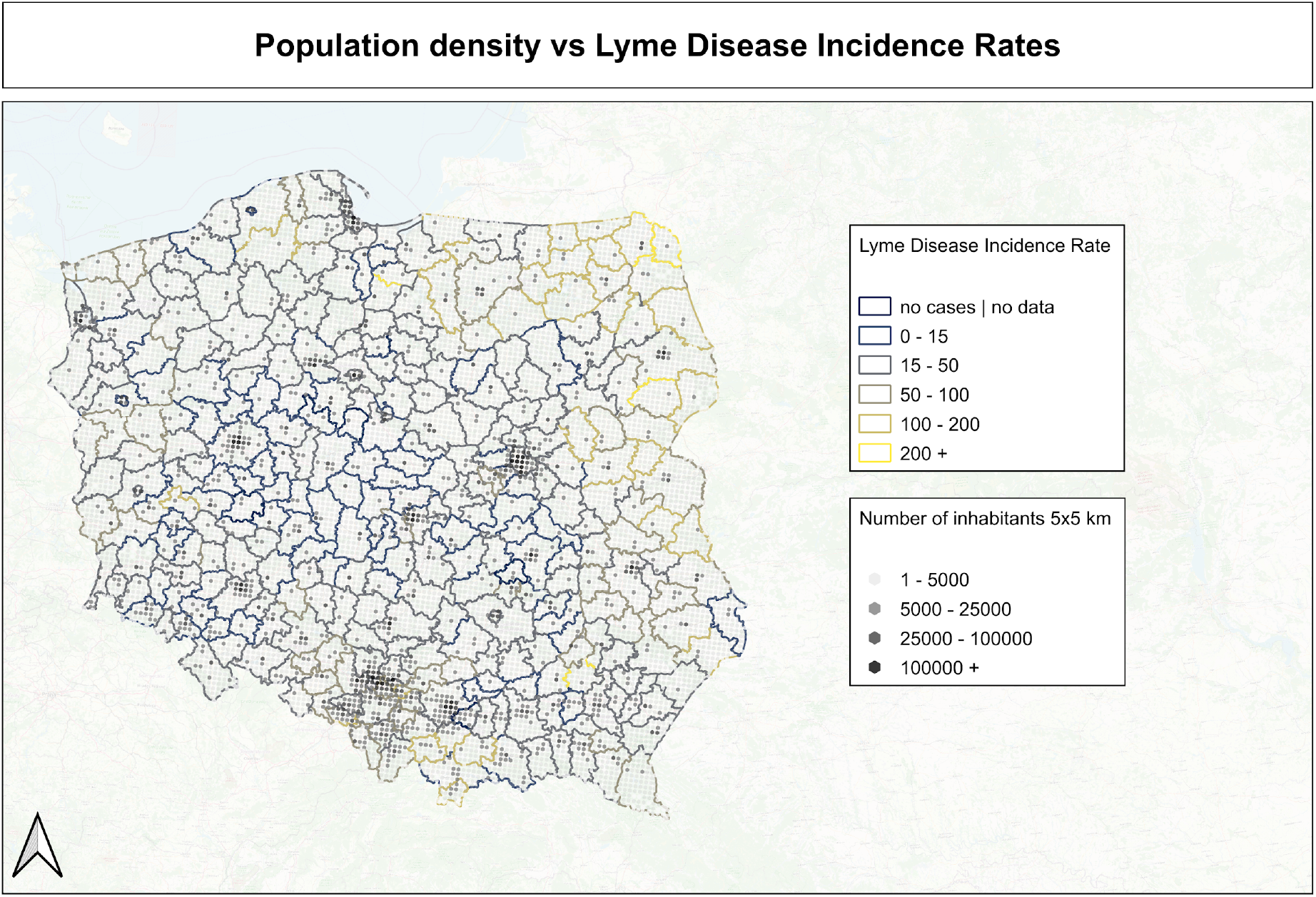
Number of inhabitants in Poland on 5×5 kilometers grid.

Population density creates point support for the deconvolution of areal incidence rate data into small and regularly spaced units. The final population-at-risk map is presented in Figure 5.

**Figure 5:**
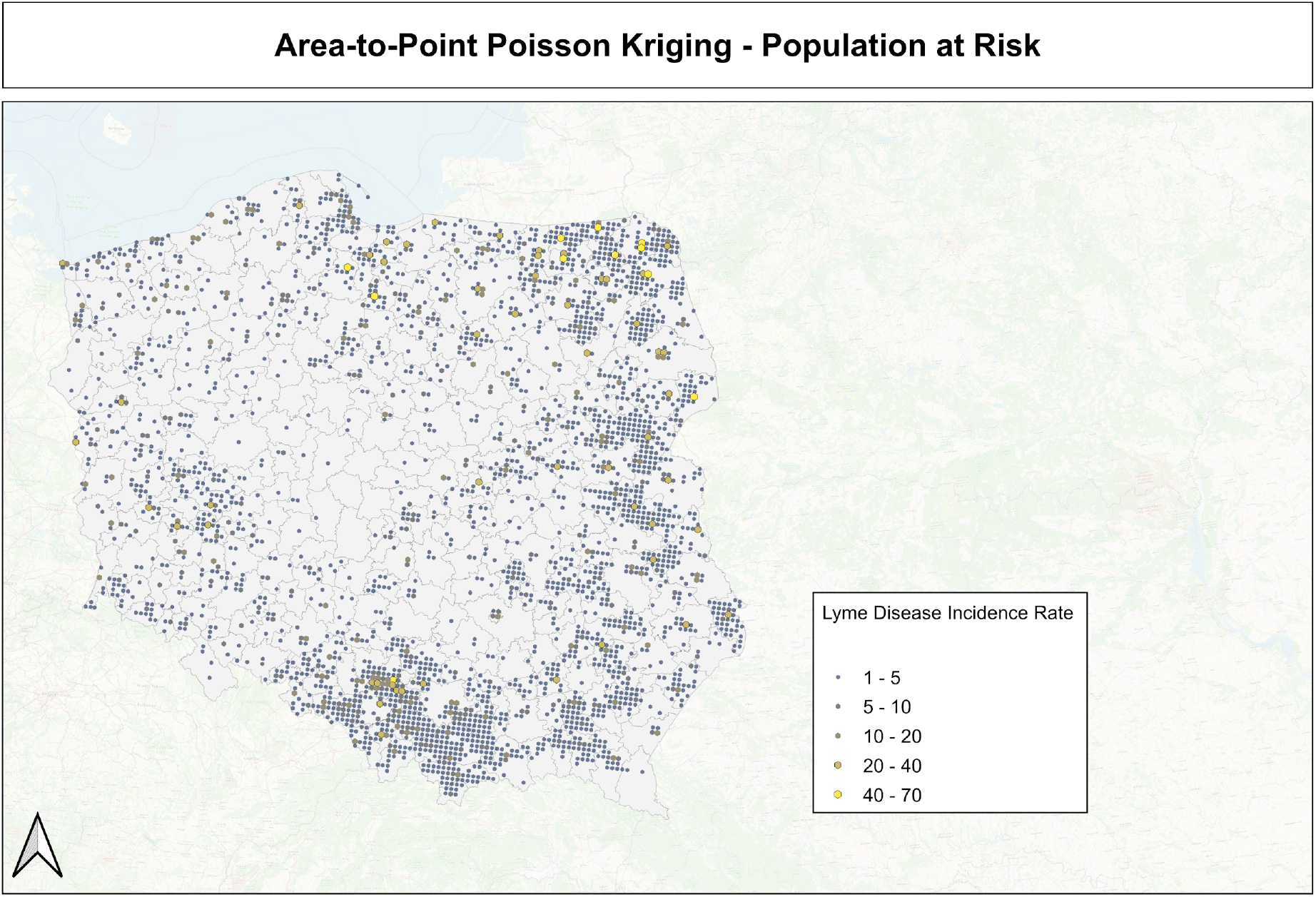
Population-at-risk after Area-to-Point Poisson Kriging transformation.

The next step is creating a species distribution map with tick occurrence probability based on the MaxEnt model and remotely sensed covariates. The map is presented in Figure 6 and shows blocks with tick occurrence probability.

**Figure 6:**
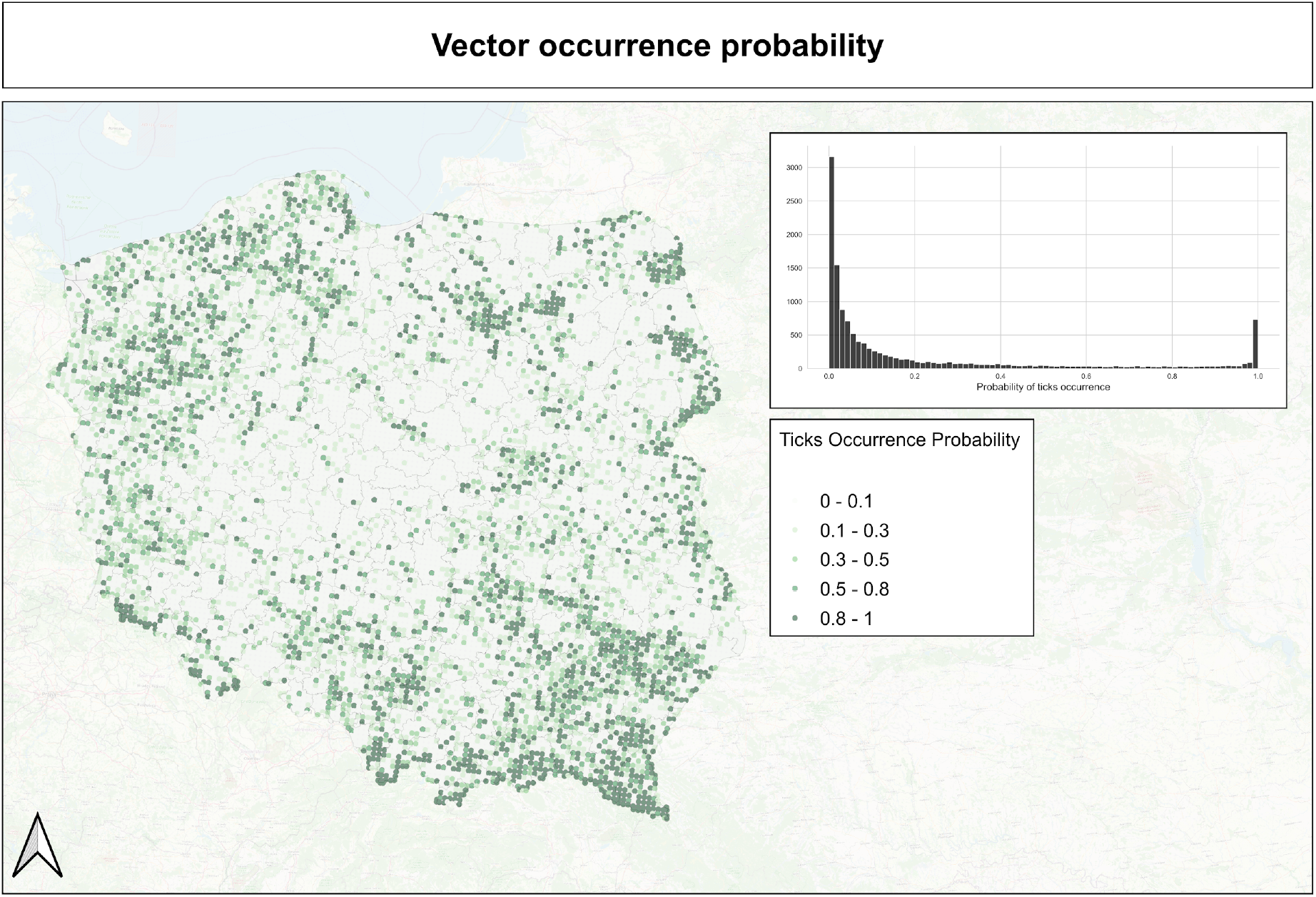
Hard-bodied ticks occurrence probability in Poland.

Both models are weighted (see equation 5), and we obtain the final weighted Lyme Disease infection risk map. Data at this stage is smoothed and ready for further analysis and decision-making (Figure 7).

**Figure 7:**
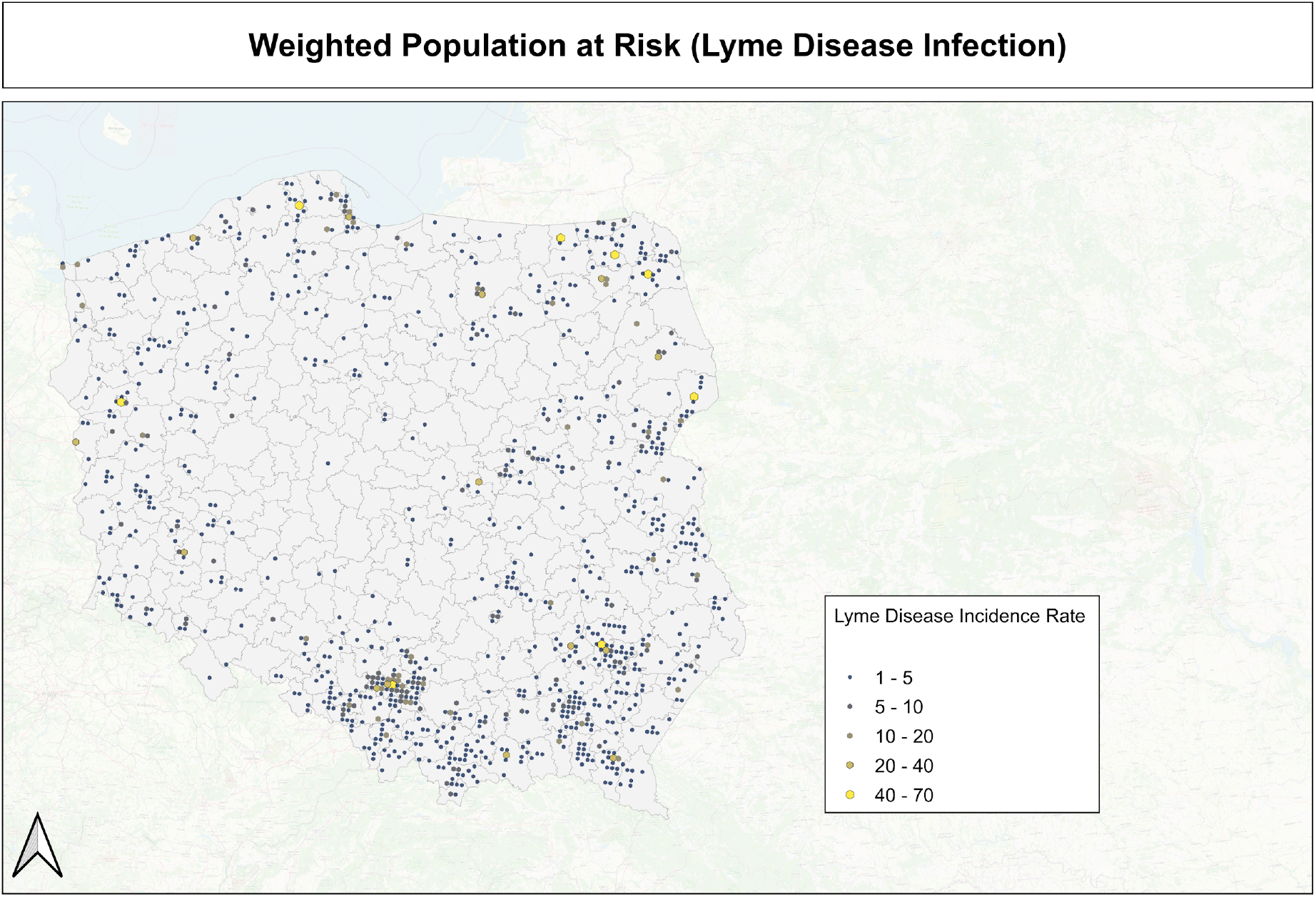
Weighted Population-at-Risk model output.

The pipeline of risk modeling is especially useful for representation of different kinds of risks. There are three outputs: population-at-risk map (1), species distribution model (2), and weighted infection-risk map (3). Every single output might be used for decision-making purposes. Figure 8 shows the approach where a researcher may focus on different aspects of disease monitoring (with filtering results that are below some risk or probability threshold).

**Figure 8:**
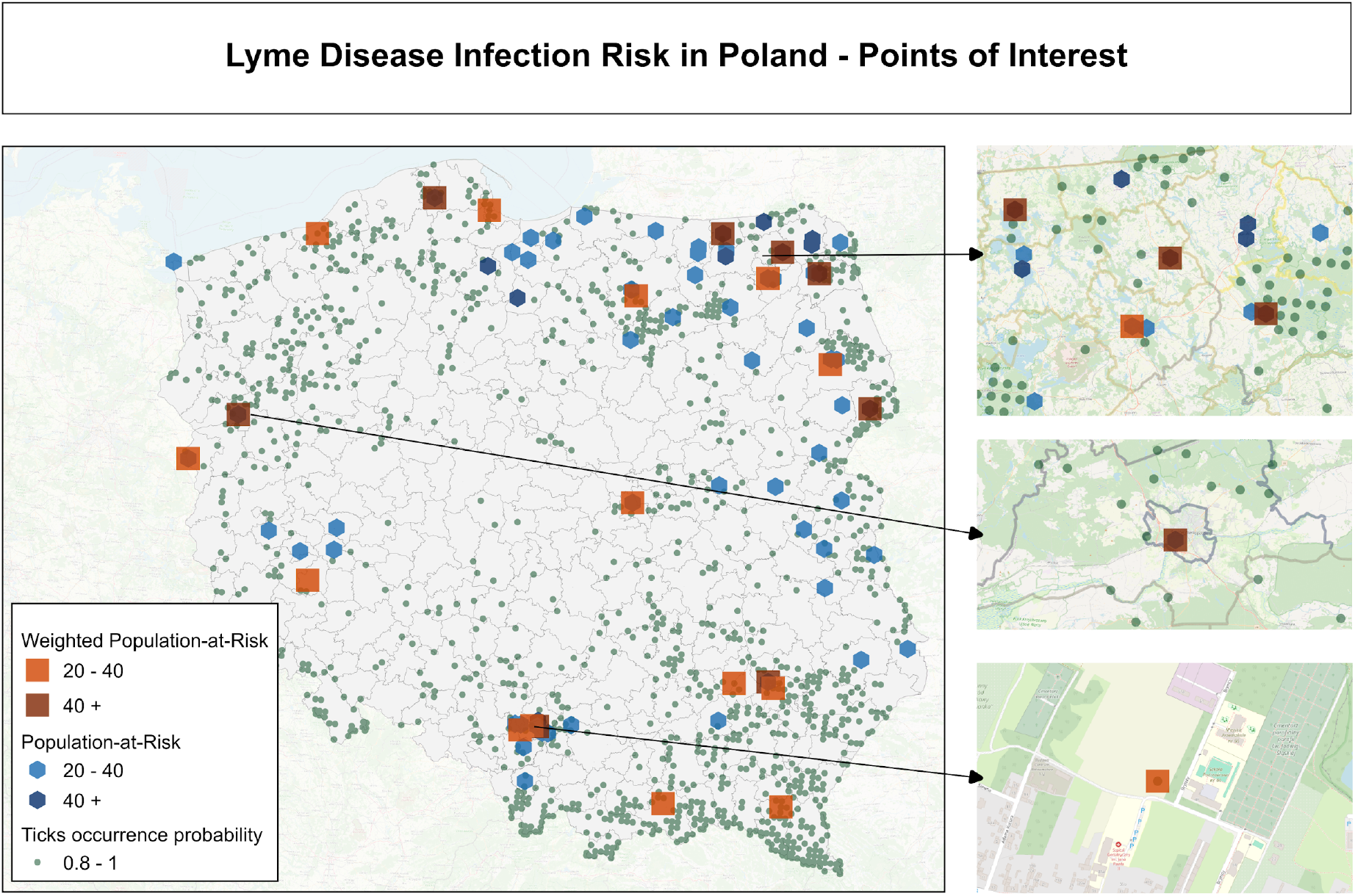
Areas that need special attention. The most dangerous regions are those where the Weighted Population at Risk is highest and are close to the environment supporting ticks’ activity.

## Discussion

The Presented vector-borne infection risk modeling pipeline is a cost-effective method. It does not require large-scale sampling or sharing of exact location data about patients. Poisson Kriging smooths aggregated incidence rates maps and transforms areal counts into population-unit blocks. The weighting step by species distribution map puts more importance on populations where the probability of vector occurrence is high, making final results more reliable for decision-making. The most important result of the analysis is when the infection risk is high and environmental factors support ticks’ existence, those areas and their population require additional monitoring.

The model has downsides. Tick bites may occur in a place different from the population block. People are traveling and hiking long distances from their homes. Thus, some infections might be “imported”, which is the model’s source of bias. The model does not estimate risk levels using weather variables that are affecting tick questing behavior, so it is only a spatial representation of the infection risk. It might be expanded into the spatio-temporal model using additional weighting factors related to weather patterns.

Using Poisson Kriging model the researcher might analyze spatial patterns and the distance at which infections are correlated. Kriging property is the generation of variance error map, which shows undersampled areas where additional sampling might be performed.

It is worth noticing that output risk map might be used as an input for complex models using high resolution remotely sensed layers and spatial information for better risk prediction. The modeling pipeline is designed for vector-borne diseases, but Poisson Kriging model alone might work with other diseases and conditions aggregated over areas as a ratio of cases per population.

## Summary

The paper presents the analytical pipeline for the interpolation of local vector-borne disease risk. The pipeline uses Area-to-Point Poisson Kriging with aggregated incidence rates and population density support for the development of the population-at-risk map and species distribution modeling for the probability of vector occurrence. The product of both components is a weighted population-at-risk map, which includes information about local infections and vector species.

## Data Availability

All data produced are available online at https://zenodo.org/records/11102795

